# Synthetic Ultrasound Image Generation for Breast Cancer Diagnosis Using cVAE-WGAN Models: An Approach Based on Generative Artificial Intelligence

**DOI:** 10.1101/2025.06.02.25328698

**Authors:** Gianluca Mondillo, Mariapia Masino, Simone Colosimo, Alessandra Perrotta, Vittoria Frattolillo, Fabio Giovanni Abbate

## Abstract

The scarcity and imbalance of medical image datasets hinder the development of robust computer-aided diagnosis (CAD) systems for breast cancer. This study explores the application of advanced generative models, based on generative artificial intelligence (GenAI), for the synthesis of digital breast ultrasound images. Using a hybrid Conditional Variational Autoencoder-Wasserstein Generative Adversarial Network (CVAE-WGAN) architecture, we developed a system to generate high-quality synthetic images conditioned on the class (malignant vs. normal/benign). These synthetic images, generated from the low-resolution BreastMNIST dataset and filtered for quality, were systematically integrated with real training data at different mixing ratios (W). The performance of a CNN classifier trained on these mixed datasets was evaluated against a baseline model trained only on real data balanced with SMOTE. The optimal integration (mixing weight W=0.25) produced a significant performance increase on the real test set: +8.17% in macro-average F1-score and +4.58% in accuracy compared to using real data alone. Analysis confirmed the originality of the generated samples. This approach offers a promising solution for overcoming data limitations in image-based breast cancer diagnostics, potentially improving the capabilities of CAD systems.

## Introduction

Breast cancer is the most common neoplasm in women [1], with projections showing increasing incidence [2]. Although early diagnosis has reduced mortality [3], the sensitivity of digital mammography (86.9% according to BCSC [4]) presents room for improvement. Computer-aided diagnosis (CAD) systems are hindered by dataset imbalance, where negative cases predominate, compromising sensitivity toward the minority pathological class. Data synthesis through generative artificial intelligence (GenAI) emerges as a solution [5], generating new samples to enrich limited datasets, balance classes, and overcome privacy restrictions. We propose a hybrid Conditional Variational Autoencoder-Wasserstein Generative Adversarial Network (CVAE-WGAN) architecture. CVAEs [6] encode images into latent distributions for controlled generation, while WGANs [7] use a stable metric for comparing distributions, improving convergence. Despite deep learning successes in mammary imaging [8], rigorous validation and interpretability remain challenges. Our objective is to demonstrate that optimized integration of generated synthetic images improves discriminative diagnostic performance in deep learning algorithms, especially for malignant lesions.

## Materials and Methods

### Dataset

We selected the BreastMNIST dataset [9], a standardized collection of low-resolution mammary ultrasound images (28×28 pixels), pre-processed by the authors. This choice was motivated by computational tractability on limited hardware. We simplified the problem to a binary classification: class 0 (malignant) vs. class 1 (normal/benign combined). The dataset was divided into training (n=491), validation (n=55), and test (n=156) sets in a percentage ratio of approximately 70% −8% −22%. The training set presents significant imbalance: 134 malignant samples (27.3%) and 357 normal/benign samples (72.7%).

### Computational Environment

Experiments conducted on: CPU: Intel Core i9-11900KF (3.50 GHz), GPU: NVIDIA RTX 4060 Ti (16GB VRAM), 32GB RAM, SSD: KIOXIA-EXCERIA G2 (1 Tb), OS: Windows 11.

### CVAE-WGAN Architecture

The architecture integrates three components: Conditional Encoder [10]: Analyzes image and class, encoding into a latent distribution (dimension 128) through residual blocks (channels 32 → 256) and multi-head attention to focus on relevant regions. The condition embedding has dimension 32. Conditional Decoder: Generates images from the latent distribution and class condition, using a structure symmetrical to the encoder. Wasserstein Discriminator [11]: Evaluates the plausibility of generated images. Uses spectral normalization and gradient penalty (*λ* = 10.0) to guarantee the Lipschitz condition and stabilize WGAN training.

### Training Procedure

Optimized training with composite loss: L1 (*γ* = 1.0), perceptual VGG (*γ* = 2.0), KL divergence (*γ* = 0.075 with annealing), Wasserstein adversarial loss (*γ* = 1.5), and feature matching (*γ* = 2.0). Adam optimizers (LR=5×10^*−*5^ for E/G/D, *β*_1_ = 0.0, *β*_2_ = 0.9) with cosine annealing scheduler and warm-up. The CVAE-WGAN architecture was trained for a total of 150 epochs, with Batch Size of 64. A Seed value of 42 was set to ensure, as much as possible with such complex architectures, maximum reproducibility of the experiment. An additional Quality Discriminator was trained (10 epochs) post-main training to filter the generated images (threshold 0.2 for malignant, 0.3 for benign). Figure 1 represents the training progress, in Figure 2 the qualitative evaluation of synthetic images generated by the model during training.

**Figure 1.**
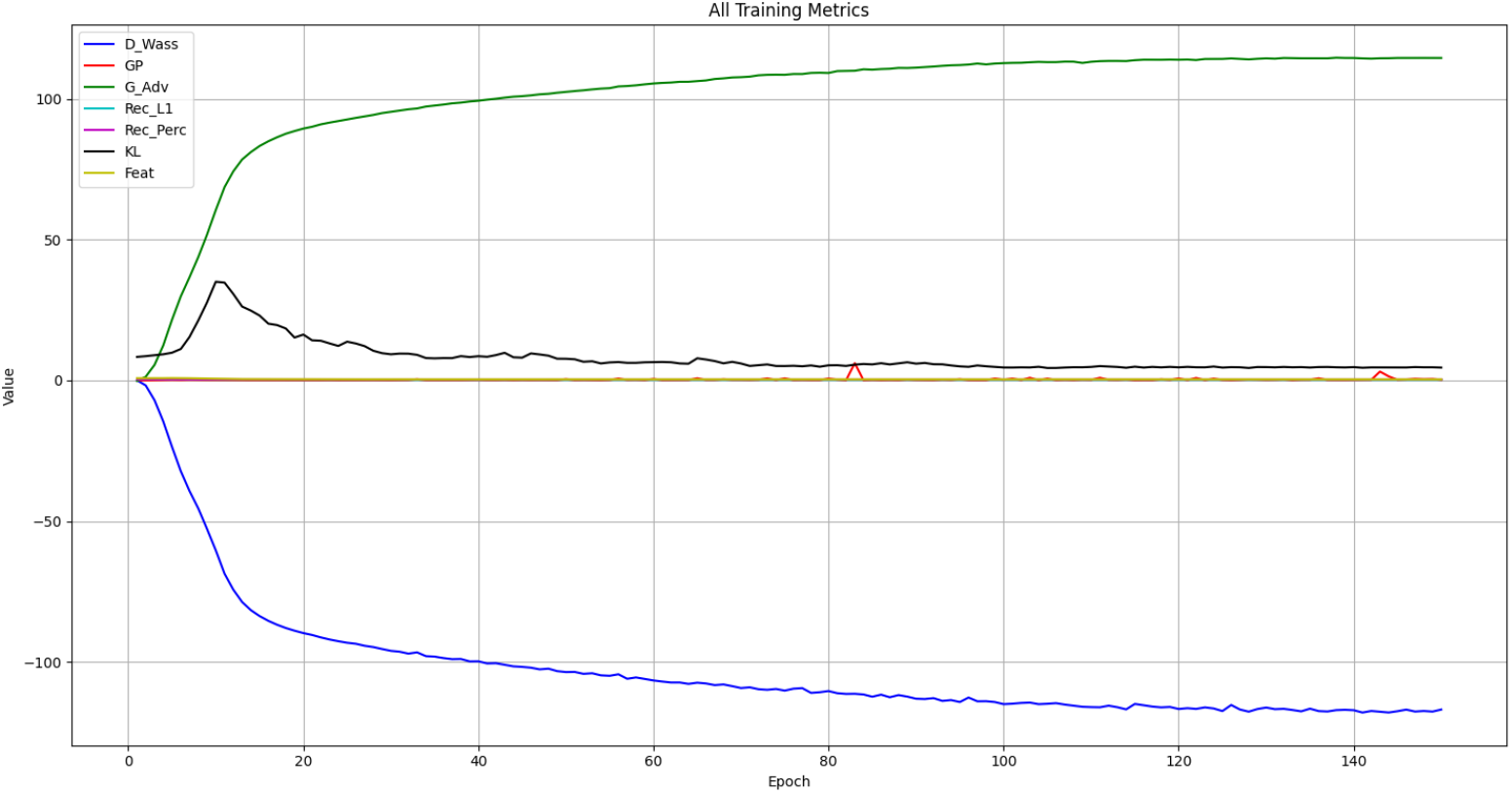
Monitoring of metrics during training of the hybrid CVAE-WGAN architecture for 150 epochs on the BreastMNIST dataset. The convergence of the discriminator’s Wasserstein loss (D Wass, blue) towards stable negative values is observed. The generator’s adversarial loss (G Adv, green) shows an increase that then stabilizes, indicating that the generator learns to produce more realistic samples. The gradient penalty (GP, red) remains close to zero, ensuring the Lipschitz condition is respected. The Kullback-Leibler divergence (KL, black) shows the expected behavior with annealing (initial peak followed by stabilization). The components related to autoencoder reconstruction (Rec L1 cyan, Rec Perc magenta) and Feature Matching loss (Feat, yellow) rapidly converge to low values, indicating good reconstruction fidelity and alignment of intermediate features between real and generated images.

**Figure 2.**
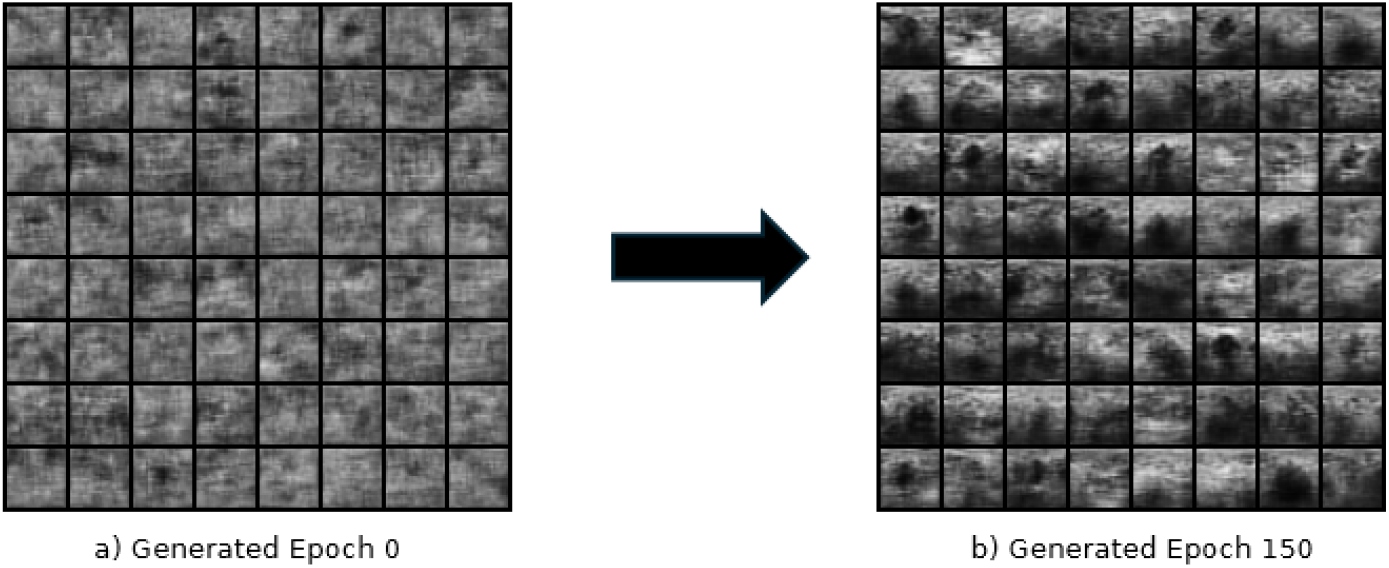
Qualitative evolution of synthetic images generated by the cVAE-WGAN model during training. On the left (a), the images produced at epoch 0 exhibit low definition and lack morphologically relevant structures. On the right (b), the images generated at epoch 150 display improved structural coherence and significantly enhanced visual quality, indicating that the model progressively learned the distributional features of the real data domain.

### Balancing and Evaluation Strategies

For the baseline (real data only), SMOTE [12] was used to balance the training set (357 samples/class). For all models, loss weights inversely proportional to class frequency were used (3.66 for malignant, 1.38 for benign). In mixed models, weighted sampling was used. The classifier’s decision threshold was calibrated by maximizing the F1-score on the precision-recall curve.

### Optimal Mix Experiment

We generated and filtered 982 synthetic samples (quality threshold arbitrarily set at 0.2). Memorization analysis showed an average Euclidean distance of 6.5093 from the closest real sample (min: 4.3526), confirming the originality of newly synthesized images. We trained classifiers: Convolutional Neural Network (CNN with 3 convolutional blocks), 2 FC, BatchNorm, Dropout on different configurations: real data only (baseline), synthetic data only, and real/synthetic mixes with relative weights (W ∈ {0.25, 0.5, 0.75, 1.0, 1.5}). Evaluation was performed on the real test set using accuracy, precision, recall, F1-score (macro-average), and AUC (macro OvR).

## Results

The FID calculated between synthetic samples and validation set is 143.34, indicating acceptable visual similarity for low-resolution medical images. Classification results are in Table 1.

**Table 1.**
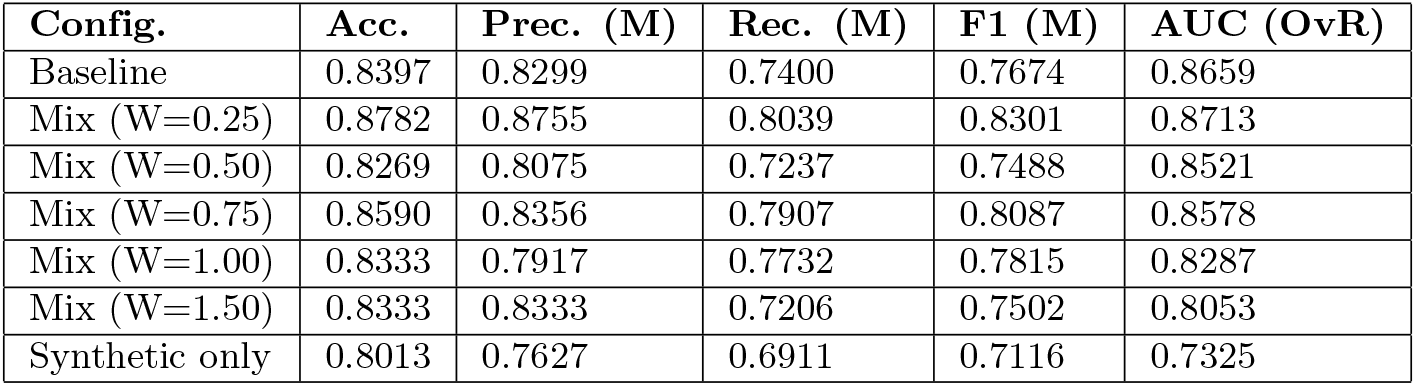
Classification results on the real test set.

| Config. | Acc. | Prec. (M) | Rec. (M) | F1 (M) | AUC (OvR) |
| --- | --- | --- | --- | --- | --- |
| Baseline | 0.8397 | 0.8299 | 0.7400 | 0.7674 | 0.8659 |
| Mix (W=0.25) | 0.8782 | 0.8755 | 0.8039 | 0.8301 | 0.8713 |
| Mix (W=0.50) | 0.8269 | 0.8075 | 0.7237 | 0.7488 | 0.8521 |
| Mix (W=0.75) | 0.8590 | 0.8356 | 0.7907 | 0.8087 | 0.8578 |
| Mix (W=1.00) | 0.8333 | 0.7917 | 0.7732 | 0.7815 | 0.8287 |
| Mix (W=1.50) | 0.8333 | 0.8333 | 0.7206 | 0.7502 | 0.8053 |
| Synthetic only | 0.8013 | 0.7627 | 0.6911 | 0.7116 | 0.7325 |

The optimal configuration (W=0.25) improved all metrics compared to the baseline. The detailed report for W=0.25 is in Table 2.

**Table 2.** Classification report for the optimal configuration (W=0.25).

| Class | Precision | Recall | F1-Score |
| --- | --- | --- | --- |
| 0 (Malignant) | 0.871 | 0.643 | 0.740 |
| 1 (Normal/Benign) | 0.880 | 0.965 | 0.921 |
| Macro Avg | 0.875 | 0.804 | 0.830 |
| Weighted Avg | 0.878 | 0.878 | 0.872 |

Compared to the baseline, the W=0.25 configuration increased accuracy by 4.58%, average precision by 5.49%, average recall by 8.64%, and average F1-score by 8.17% (AUC +0.63%). To further highlight the impact of synthetic data augmentation on class-specific performance, Table 3 presents a direct comparison of per-class classification metrics between the baseline configuration (real data augmented with SMOTE during classifier training) and the optimal W=0.25 configuration (real data augmented with GAN-generated synthetic data). Figure 3 shows a representation of conditional images from BreastMNIST vs Data Synthesis.

**Table 3.** Per-class classification metrics comparison for Baseline vs. Optimal Mix (W=0.25) on the real test set.

| Configuration | Class | Precision | Recall | F1-Score |
| --- | --- | --- | --- | --- |
| Baseline | 0 (Malignant) | 0.815 | 0.524 | 0.638 |
|  | 1 (Normal/Benign) | 0.845 | 0.956 | 0.897 |
| Mix ($W=0.25$) | 0 (Malignant) | 0.871 | 0.643 | 0.740 |
|  | 1 (Normal/Benign) | 0.880 | 0.965 | 0.921 |

**Figure 3.**
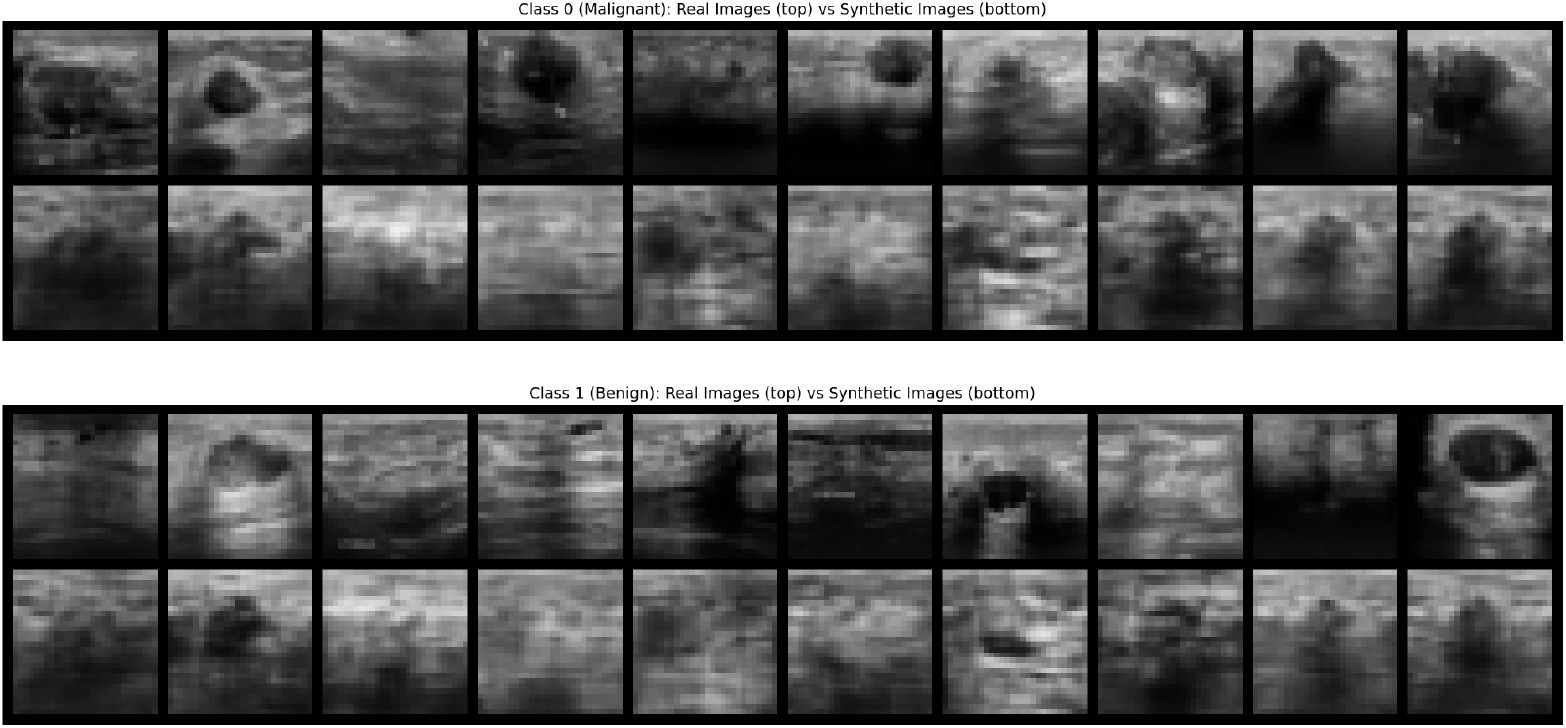
Visual comparison between real images (upper row of each panel) and synthetic images (lower row) generated by the cVAE-WGAN. Upper panel: Class 0 (Malignant). Lower panel: Class 1 (Normal/Benign).

As shown in Table 3, the W=0.25 configuration substantially improved the recognition of the minority ‘0 (Malignant)’ class, with its Recall increasing from 0.524 to 0.643 and its F1-Score improving from 0.638 to 0.740. This demonstrates the cVAE-WGAN’s effectiveness in augmenting the dataset for the underrepresented class, leading to a more balanced and robust classifier. The t-SNE analysis of the latent space (perplexity=30.0, n=55) showed well-separated class clusters (Figure 4).

**Figure 4.**
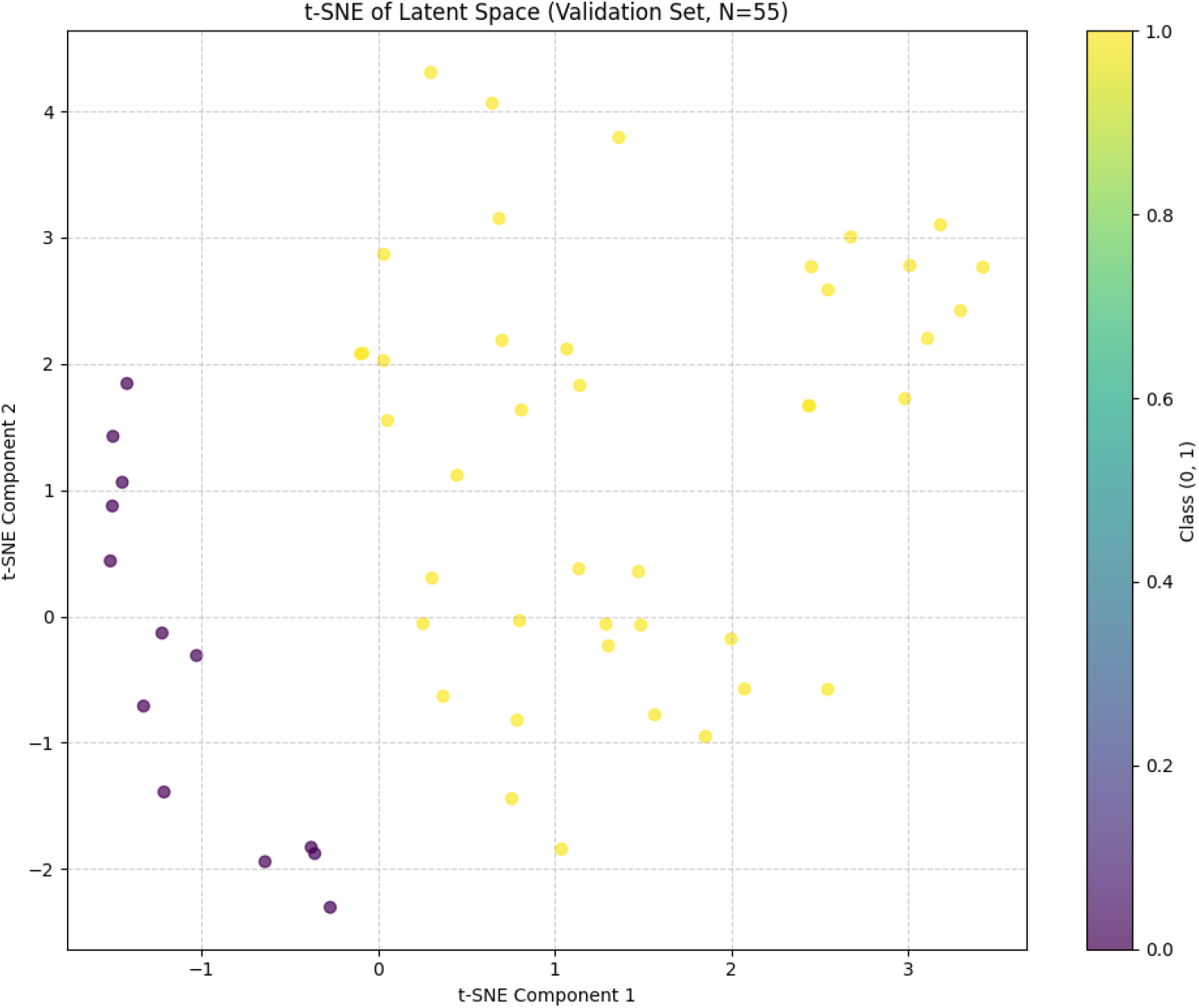
Two-dimensional t-SNE projection of the latent space (dimension 128) generated by the CVAE-WGAN model’s encoder applied to N=55 real samples from the validation set. Colors indicate the real class (Class 0: Malignant; Class 1: Normal/Benign), as shown in the color bar. The clear separation of clusters suggests that the model has learned a discriminative latent representation of real data. A well-structured and class-separated latent space is fundamental for controlled image generation. It means that by sampling from different regions of the latent space and providing the appropriate label to the decoder, there is a higher probability of generating synthetic images that resemble the desired class.

## Discussion

The optimal integration of synthetic data generated via a CVAE-WGAN architecture demonstrated a significant improvement in diagnostic performance, with an 8.17% increase in F1-score that is clinically relevant. Of particular clinical significance is the 11.9 percentage point increase in recall for the malignant class achieved by our optimal configuration (Table 3). Improving the sensitivity for detecting malignancy is paramount in oncologic diagnostics, as it directly translates to a reduced rate of false negatives, potentially leading to earlier treatment and better patient outcomes. This result is consistent with recent evidence confirming the effectiveness of synthetic images for medical dataset augmentation [5]. Our optimal mixing ratio (W=0.25) is supported by the studies of Yu et al. [15], who showed that integrating synthetic and real data outperforms the use of real data alone, and that models trained exclusively on synthetic data can approach the performance of those trained on real data. This is particularly relevant considering that the model trained only on synthetic data in our study achieved an accuracy of 80.13% and an F1-score of 71.16%, demonstrating the potential value of synthetic data in data-scarce scenarios. The use of SMOTE to rebalance the baseline represents a methodologically sound strategy to ensure fair comparisons, avoiding bias introduced by dataset imbalance. This approach aligns with the best practices indicated by Kebaili et al. [5], who emphasize the importance of appropriate augmentation techniques in image-based diagnostics. Our CVAE-WGAN approach differs significantly from prior literature due to several innovative features. The integration of CVAE and WGAN represents an advancement over single-model approaches. Zama et al. [16] demonstrated the effectiveness of DCGANs in generating synthetic breast ultrasound images, achieving promising results but without incorporating the conditional generation capabilities of CVAEs. Our hybrid architecture combines the training stability of WGANs with the controllability of conditional generation offered by CVAEs. The implementation of an additional quality discriminator with class-specific thresholds (0.2 for malignant, 0.3 for benign) represents a methodological innovation not reported in previous studies [13–15], enabling an automated filtering of generated images and ensuring high quality without manual intervention. The systematic evaluation of different mixing ratios (W ∈ {0.25, 0.5, 0.75, 1.0, 1.5}) provides quantitative insights into the optimal integration of synthetic and real data. This analysis goes beyond previous studies that often relied on qualitative assessments or fixed ratios [11, 14]. The FID score of 143.34 obtained in our study, although higher than those reported for high-resolution datasets [11], is considered acceptable in the context of low-resolution medical images (28×28 pixels). Müller-Franzes et al. [17] have shown that diffusion models can outperform GANs in terms of diversity and quality, but require significantly greater computational resources. The memorization analysis, with a mean Euclidean distance of 6.5093 from the closest real sample, confirms the generation of original samples and counters the memorization bias commonly associated with generative models, as highlighted by Khader et al. [18] in their work on diffusion models for 3D images. The 8.17% improvement in F1-score significantly exceeds previous literature results: Shen et al. [14] reported gains of up to 5.03%, while studies focusing on visual validation [13, 14, 16] did not provide comparable quantitative metrics. Our increases in accuracy (4.58%) and recall (8.64%) further demonstrate the effectiveness of the proposed approach. The study by Jiménez-Gaona et al. [11] emphasized the importance of tailoring GAN architectures to specific modalities (SNGAN for mammography, cGAN for ultrasound). Our CVAE-WGAN architecture represents an evolution that aims to combine the strengths of various frameworks, offering greater versatility compared to highly specialized models. The main limitation of our study concerns the low resolution (28×28 pixels) of the BreastMNIST dataset. This choice, driven by computational feasibility, compromises the capture of fine details that are crucial for clinical diagnostics. Alruily et al. [19] demonstrated that high-resolution ultrasound image augmentation requires more sophisticated architectures, such as GANs with modified identity blocks. Binary classification (malignant vs. normal/benign) represents a simplification compared to the complexity of real-world diagnostic scenarios. The absence of radiological validation due to the low resolution constitutes a significant limitation. Zama et al. [16] conducted reader studies with expert radiologists, showing that synthetic DCGAN images can convincingly mimic real clinical images. Such clinical validation will be essential for future iterations of our method. The use of synthetic images raises important ethical and privacy concerns. Schreiner et al. [20] demonstrated that GANs can overcome barriers to data sharing by generating fully synthetic datasets that preserve aggregate statistical properties without containing identifiable patient information. However, as highlighted by Müller-Franzes et al. [17], the risk of training data memorization persists. Our Euclidean distance analysis (mean 6.5093) provides preliminary evidence against direct memorization, but future studies should implement more sophisticated metrics to ensure patient privacy. Future implementations should focus on extending to higher resolutions. Recent developments have demonstrated the feasibility of scalable approaches for generating high-resolution medical images. Combining techniques such as progressive growing and multi-scale discriminators may allow the adaptation of our CVAE-WGAN architecture to clinically relevant resolutions. The integration of multiple validation modalities, including automated quantitative assessments and reader studies with radiologists, is an essential step toward clinical translation. Allan-Blitz et al. [21] demonstrated that the synergistic pairing of synthetic image generation with pathological classification models enables the rapid development of digital diagnostic tools. The lack of standardized metrics for evaluating synthetic medical images remains a significant challenge. Baltruschat et al. [22] identified five major pitfalls in the use of reference metrics for synthetic medical images, stressing the need to develop domain-specific metrics. The proposed CVAE-WGAN model demonstrates effectiveness in augmenting datasets for breast ultrasound diagnosis, with measurable improvements in classification performance. This approach represents a significant step toward the systematic use of GenAI in oncologic diagnostics, from algorithm training to clinical education. However, clinical translation will require further development in terms of resolution, multidisciplinary validation, and standardization of evaluation metrics. Ethical and privacy considerations must guide future developments, ensuring that the benefits of synthetic augmentation do not compromise patient safety and privacy. Emerging evidence suggests that the optimal integration of synthetic and real data, rather than complete substitution, represents the most promising strategy for improving diagnostic performance. The identified optimal mixing ratio (W=0.25) provides an empirical starting point for future implementations, although validation in diverse clinical settings is still required.

## Conclusions

CVAE-WGAN models are promising for mitigating data scarcity and imbalance in breast cancer imaging. The optimal integration (W=0.25) of synthetic data significantly improves diagnostic performance (F1-score +8.17%, Acc +4.58%). Latent space analysis confirms the learning of meaningful representations. Future perspectives include application to high-resolution datasets, multiclass models, clinical validation, and exploration of advanced techniques for diagnostic personalization. The proposed approach supports the use of GenAI in oncological diagnostics, from algorithm training to clinical education.

## Data Availability

All data produced in the present study are available upon reasonable request to the authors.

